# Gender Discrimination and Medical Student Development

**DOI:** 10.1101/2025.02.06.25321787

**Authors:** Shruthi Venkataraman, Mytien Nguyen, Sarwat I. Chaudhry, Mayur M. Desai, Tonya L. Fancher, Alexandra M. Hajduk, Hyacinth R. C. Mason, Alexis Webber, Dowin Boatright

## Abstract

**Background:** Despite prevalent gender discrimination in medical education, its influence on personal and professional development, foundational competencies in medical training per the Association of American Medical Colleges (AAMC), remains unclear. This retrospective cross-sectional study assesses how experiences of gender discrimination in medical school influence personal and professional identity formation (PPIF).

**Methods:** Deidentified student-level data were procured from the AAMC data warehouse for 37,610 MD students who matriculated in 2014-2015 and took the Graduation Questionnaire (GQ) between 2016-2020. Gender discrimination frequency was categorized as ‘Never’, ‘Isolated’, and ‘Recurrent’ from GQ responses to questions about denial of opportunities, offensive remarks, and lower evaluations due to gender. Gender was binarized, due to dataset limitations. PPIF was assessed using two GQ metrics, personal and professional development, and dichotomized.

**Results:** Female students experienced higher rates of isolated (12.6%) and recurrent (20.1%) gender discrimination than males (4.3% isolated, 6.2% recurrent). Females reported slightly lower personal (71.2%) but similar professional development (92.2%) rates compared to males (73.4% personal, 91.2% professional). Both genders experiencing gender discrimination had lower likelihoods of PPIF than their counterparts without these experiences. If recurrent discrimination occurred, the aRR (95%CI) of professional development was 0.89 (0.87-0.90) for females and 0.78 (0.74-0.81) for males, while for personal development, it was 0.69 (0.67-0.71) for females and 0.61 (0.58-0.66) for males. Compared to females, males showed sharper declines in professional development as discrimination frequency increased from never to isolated (exp(b)=0.93, 95% CI [0.92-0.94], p<0.001) and isolated to recurrent (exp(b)=0.95, 95% CI [0.93-0.97], p<0.001).

**Conclusions:** Gender discrimination negatively influences PPIF for both female and male medical students. Efforts to combat discrimination in medical training and promote holistic student development should be considered. Future work is needed to understand the influence of gender discrimination on the comprehensive development of gender-diverse medical students.

## Introduction

The Association of American Medical Colleges (AAMC) emphasizes the importance of personal and professional development—cultivating qualities necessary for lifelong growth as a person and a physician—as foundational competencies in medical training.(1) The learning environment in medical school shapes identity formation and is influenced by interactions with peers, patients, and supervisors.(2–4)

However, the medical training environment is not exempt from the broader social challenge of gender discrimination.(5) In a 1995 report, the Council on Graduate Medical Education stated: “Gender bias, a reflection of society’s value system, remains the single greatest deterrent to women achieving their full potential in every aspect of the medical profession and is a barrier throughout the professional life cycle.”(6)

Research consistently shows that female students face more gender-based discrimination and harassment,(7–9) but males are not entirely spared. In specialties like obstetrics and gynecology and pediatrics, male students report facing discrimination, especially regarding mentoring, access to educational opportunities, and encouragement to pursue these fields.(6)

Despite the prevalence of gender discrimination in medical education,(10) its impact on the ability of medical schools to foster personal and professional identity formation (PPIF) among students remains unclear. This study investigates differences in the influence of gender discrimination on PPIF among men and women in medical school.

## Methods

### Data and Participants

Using fully anonymized student-level data from the AAMC data warehouse, we conducted a retrospective cross-sectional study of 40,971 Doctor of Medicine (MD) matriculants from the 2014-2015 and 2015-2016 academic years who graduated between 2016 and 2020 and took the American Medical College Application Service (AMCAS) survey and Graduation Questionnaire (GQ). Students provided explicit electronic consent on AMCAS by actively selecting an option to allow the use of their data for research purposes. Participation in the GQ was voluntary, and students were informed that their responses could be used for research purposes, providing implied informed consent by choosing to participate. The Yale School of Medicine institutional review board waived the need for additional ethics review and informed consent, as all data were fully deidentified by the AAMC prior to transfer and researcher access. Data were accessed for research purposes between February 1, 2024, and June 3, 2024. The study followed the Strengthening the Reporting of Observational Studies in Epidemiology reporting guidelines for cross-sectional studies. Four students with unknown gender and 3,357 students who did not graduate from medical school were not included in the analysis. The final sample included 37,610 participants.

### Student Sociodemographic Characteristics

Data on gender, race and ethnicity, and childhood household income were obtained from the AMCAS. Students self-reported their gender as male or female. Students self-reported their race and ethnicity as corresponding to any or all the following groups: African American or Black, American Indian or Alaska Native (AIAN), Asian, Hawaiian Native or Pacific Islander (HNPI), Hispanic, Latino, or of Spanish Origin, white, other, and unknown. Students reporting identification with 2 or more groups were categorized as multiracial. Students self-selected their childhood household income from 17 categories, which were then recategorized into 5 ranges to approximate income quintiles: less than $50,000, $50,000 to less than $75,000, $75,000 to less than $125,000, $125,000 to less than $200,000 and $200,000 or more.

### Experience of Gender Discrimination

Gender discrimination frequency was determined via the following three questions in the GQ: 1) “During medical school, how frequently have you been denied opportunities for training or rewards based on gender?” 2) “During medical school, how frequently have you been subjected to sexist remarks/names?” and 3) “During medical school, how frequently have you received lower evaluations or grades solely because on gender rather than performance?” To each question, participants could respond ‘Never’, ‘Once’, ‘Occasionally’ or ‘Frequently’. We recategorized the variable into three levels: none, isolated, and recurrent. Participants who reported ‘None’ to all three questions were categorized as having no experiences of gender discrimination. Those who responded ‘Once’ to only one of the questions were categorized as having isolated experiences. Participants were classified as having recurrent experiences if they met one of the following criteria: at least one response of ‘Occasionally’ or ‘Frequently’ to any question, or ‘Once’ to more than one question.

### Outcomes

PPIF was assessed using two distinct metrics: personal and professional development. Personal development was measured by the GQ item “My medical school has done a good job of fostering and nurturing my development as a person” and professional development was measured by the GQ item “My medical school has done a good job of fostering and nurturing my development as a physician”. Responses were recorded on a 5-point Likert scale from ‘Strongly disagree’ to ‘Strongly agree’. Variables were dichotomized such that only participants who expressed strong agreement (i.e., ‘Agree’ or ‘Strongly agree’) were considered as reporting that their medical school nurtured their PPIF. Both personal and professional development were examined separately in all analyses; the term PPIF is used to collectively refer to these two outcomes.

### Statistical Analysis

All data analyses were conducted from December 1, 2023, to July 12, 2024, using Stata version 16.1 (StataCorp LLC). Missing data were present across several key variables, with the extent of missingness varying. While there were no missing data for binary gender, 6.46% of the data were missing for race/ethnicity, 22.43% of the data were missing for childhood household income, 27.5% of data were missing for experience of gender discrimination, 20.6% for personal development, and 19.8% for professional development. A total of 21,728 participants answered all the questionnaire items relevant to this study.

Missing data were imputed using a fully conditional specification method to handle arbitrary missing patterns across all categorical data. The imputation model included all sociodemographic variables, experience of gender discrimination and PPIF outcome variables. Twenty imputed data sets were created. To investigate the independent effect of gender on the PPIF outcomes, we utilized generalized linear models (GLMs) with a Poisson distribution and robust standard errors wherein gender or experience of gender discrimination were independent variables and personal and professional development were the outcome variables. We used this method because PPIF outcomes were common.(11) To assess whether experience of gender discrimination moderates the influence of binary gender on PPIF, we stratified men and women participants based on the level of gender discrimination experience and conducted GLMs with a Poisson distribution and robust standard errors. To test whether the changes in adjusted relative risk of PPIF with increasing levels of gender discrimination differ by gender, we ran the regression models with an interaction term. Sensitivity analysis considering only complete cases yielded similar findings, and multiple imputation resulted in a better fitting model, so only the post-imputation results are presented. Two-sided *p* < .05 indicated significance. All relative risk estimates are adjusted for race, ethnicity and childhood household income.

## Results

The imputed dataset (N=37,610) showed a slightly lower proportion of females (48.4%) than males (***Table***). Over 20% of participants reported experiencing gender discrimination, with 12.9% experiencing it recurrently. Most students reported that their medical schools fostered their personal (72.3%) and professional (91.7%) development, with a notably higher endorsement of professional development.

### Prevalence of Gender Discrimination among Female and Male Students

Among females, 20.1% (N=3658) experienced recurrent, 12.6% (N=2293) experienced isolated, and 67.3% (N=12,249) never experienced gender discrimination (***Figure 1***). Among males, 6.2% (N=1203) experienced recurrent, 4.3% (N=835) experienced isolated, and 89.5% (N=17,372) never experienced gender discrimination. Experiences of gender discrimination were significantly more common among female students compared to their male counterparts (χ^2^(2)=2769.4, p < 0.001).

**Figure 1:**
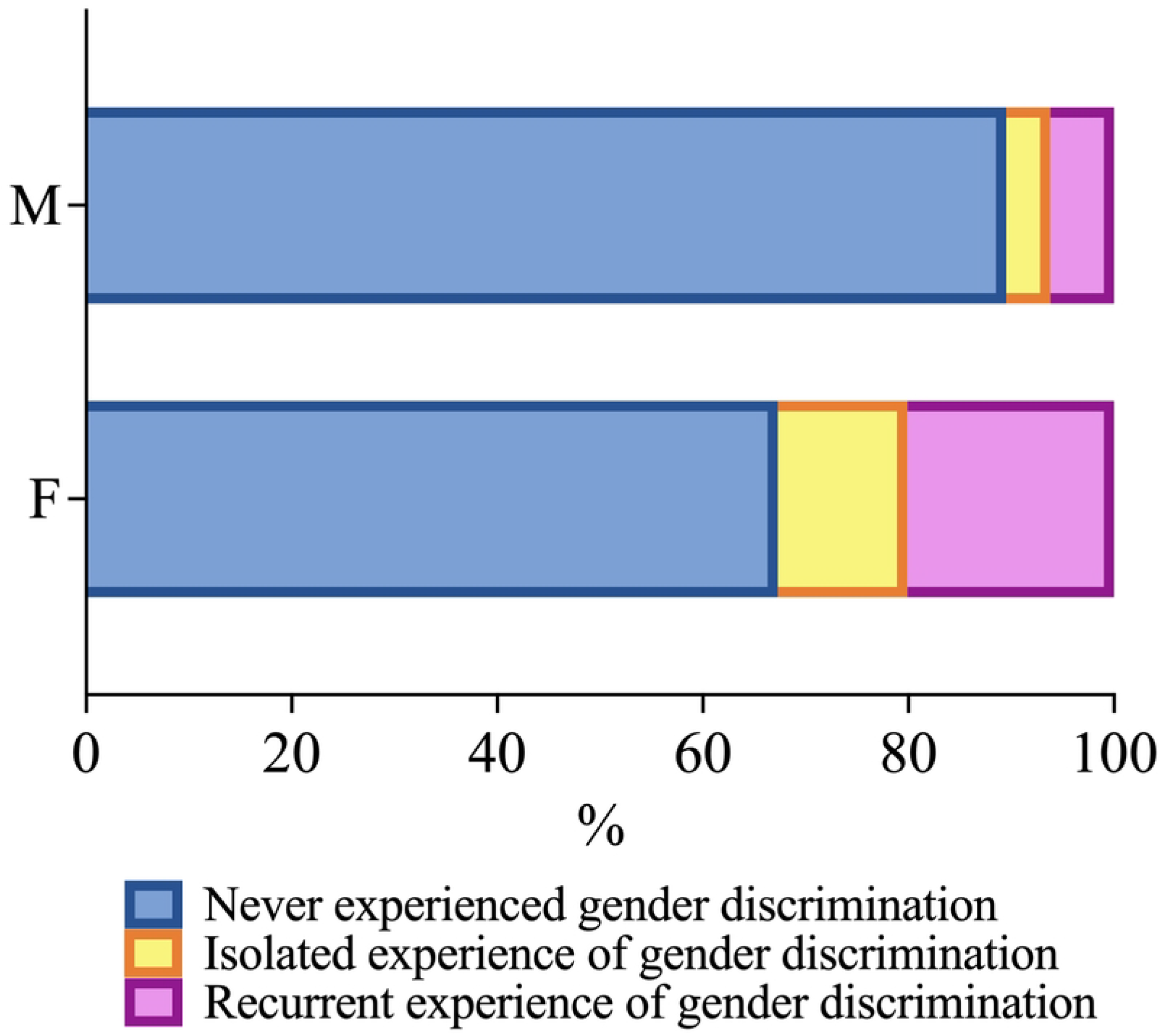
Frequency of gender discrimination experienced by male (M) and female (F) medical students. The stacked bar charts display the percentages of male and female medical students who experienced gender discrimination. The data is categorized into three groups: those who never experienced gender discrimination (blue), those who had isolated experiences of gender discrimination (yellow), and those who had recurrent experiences of gender discrimination (pink).

### Influence of Binary Gender on PPIF in Medical Education

Female students were less likely to report that their medical school fostered and nurtured their personal development (71.2%, N=12,950) than male students (73.4%, N=14,246, χ^2^(1)=23.57, p<0.001, ***Figure 2A***). Compared to males, females reported a lower likelihood of their medical school fostering their personal development (adjusted Relative Risk; aRR=0.97, 95% CI 0.96-0.98). Female students were more likely to report that their medical school fostered and nurtured their professional development (92.2%, N=17,705) than male students (91.2%, N=16,796, χ^2^(1)=1430.75, p<0.001, ***Figure 2B***). However, the likelihood of agreeing that their medical school fostered their professional development was not significantly different for females compared to males after adjustment for race/ethnicity and childhood household income (aRR=1.01, 95% CI 1.00-1.02).

**Figure 2:**
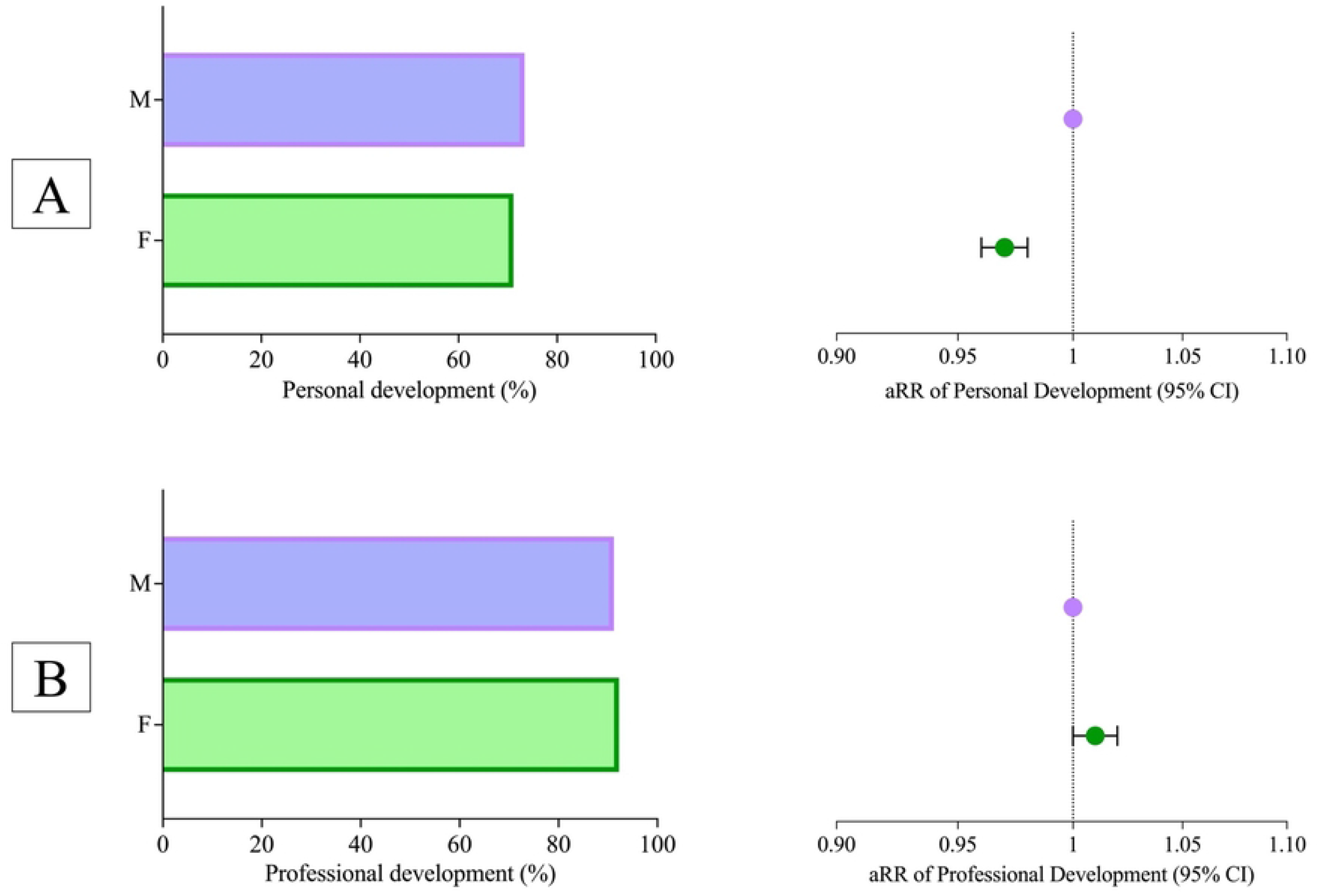
Personal and Professional Development Among Medical Students by Gender. Panel A shows the proportion of female (F) and male (M) medical students reporting that their medical school fostered their personal development. The associated forest plot on the right illustrates the adjusted relative risk (aRR) for personal development with 95% confidence intervals, comparing female students to their male counterparts (reference group). Panel B shows the proportion of female and male medical students reporting that their medical school fostered their professional development. The associated forest plot on the right illustrates the aRR for professional development with 95% confidence intervals, comparing female students to their male counterparts. All relative risks were adjusted for race/ethnicity and childhood income.

### Influence of Gender Discrimination on PPIF in Medical Education among Female and Male Students

Among females, the prevalence and likelihood of students reporting that their medical school fostered their personal development was inversely proportional to the frequency of gender discrimination: 77.2% (N=9,443, reference for aRR) if none, 67.3% (N=1,549, aRR=0.87, 95%CI 0.84-0.90) if isolated, and 53.3% (N=1,957, aRR=0.69, 95%CI 0.67-0.71) if recurrent (χ^2^(2)=788.37, p<0.001, ***Figure 3A***). Patterns for professional development were similar: 94.5% (N=11,565, reference for aRR) if none, 92.8% (N=2,138, aRR=0.98, 95%CI 0.96-0.99) if isolated, and 84.2% (N=3,094, aRR=0.89, 95%CI 0.87-0.90) if recurrent (χ^2^(2)=397.19, p<0.001, ***Figure 3B***).

**Figure 3:**
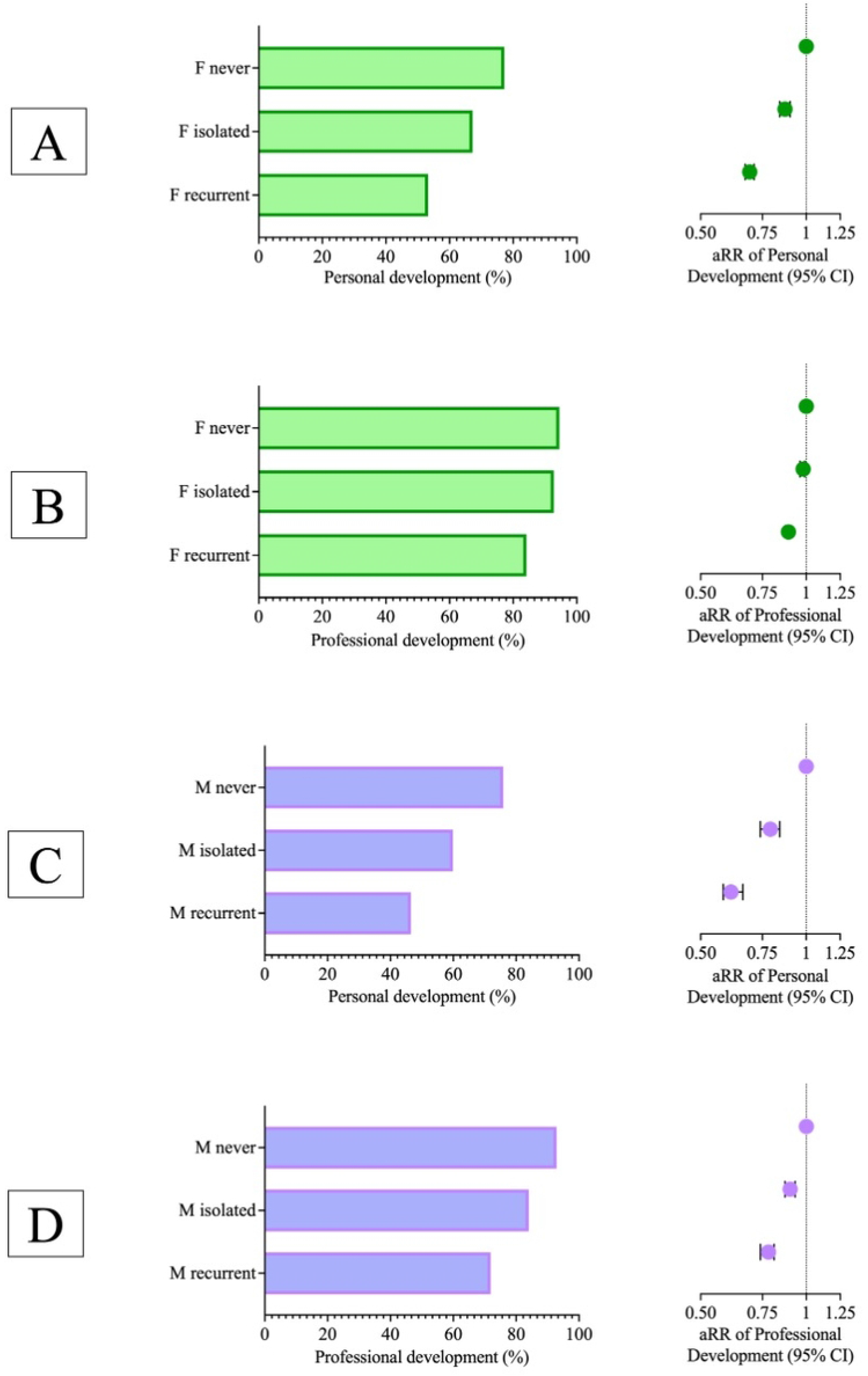
Personal and Professional Development Among Female and Male Medical Students by Frequency of Gender Discrimination. Panels A and C show horizontal bar charts depicting the percentages of personal development for females and males who never experienced, had isolated experiences of, or recurrent experiences of gender discrimination, respectively. The associated forest plots on the right presents the adjusted relative risk (aRR) of personal development for females or males with 95% confidence intervals, based on the level of gender discrimination experienced. Panels B and D show horizontal bar charts depicting the percentages of professional development for males who never experienced, had isolated experiences of, or recurrent experiences of gender discrimination, respectively. The associated forest plots on the right presents the aRR of professional development for females or males with 95% confidence intervals, according to the level of gender discrimination experienced. All relative risks were adjusted for race/ethnicity and childhood income.

Among males, the prevalence and likelihood of students reporting that their medical school fostered their personal development was inversely proportional to the frequency of gender discrimination: 75.9% (N=13,185, reference for aRR) if none, 59.9% (N=504, aRR=0.79, 95%CI 0.74-0.84) if isolated, and 46.5% (N=557, aRR=0.61, 95%CI 0.58-0.66) if recurrent (χ^2^(2)=565.83, p<0.001, ***Figure 3C***). Patterns for professional development were similar: 92.9% (N=16,145, reference for aRR) if none, 84% (N=703, aRR= 0.90, 95%CI 0.87-0.93) if isolated, and 71.9% (N=857, aRR=0.78, 95%CI 0.74-0.81) if recurrent (χ^2^(2)=680.91, p<0.001, ***Figure 3D***).

### Sharper Declines in PPIF Rates for Males with Increasing Frequency of Gender Discrimination

The change in predicted relative risk of the school fostering personal development among students who never experienced gender discrimination compared to students who experienced isolated discrimination was significantly higher in male students than female students (exp(b)=0.90, 95% CI 0.88-0.92, p<0.001, ***Figure 4A***). The change in predicted relative risk of the school fostering personal development among students who experienced isolated gender discrimination compared to those who experienced recurrent discrimination was not significantly different between both genders.

**Figure 4:**
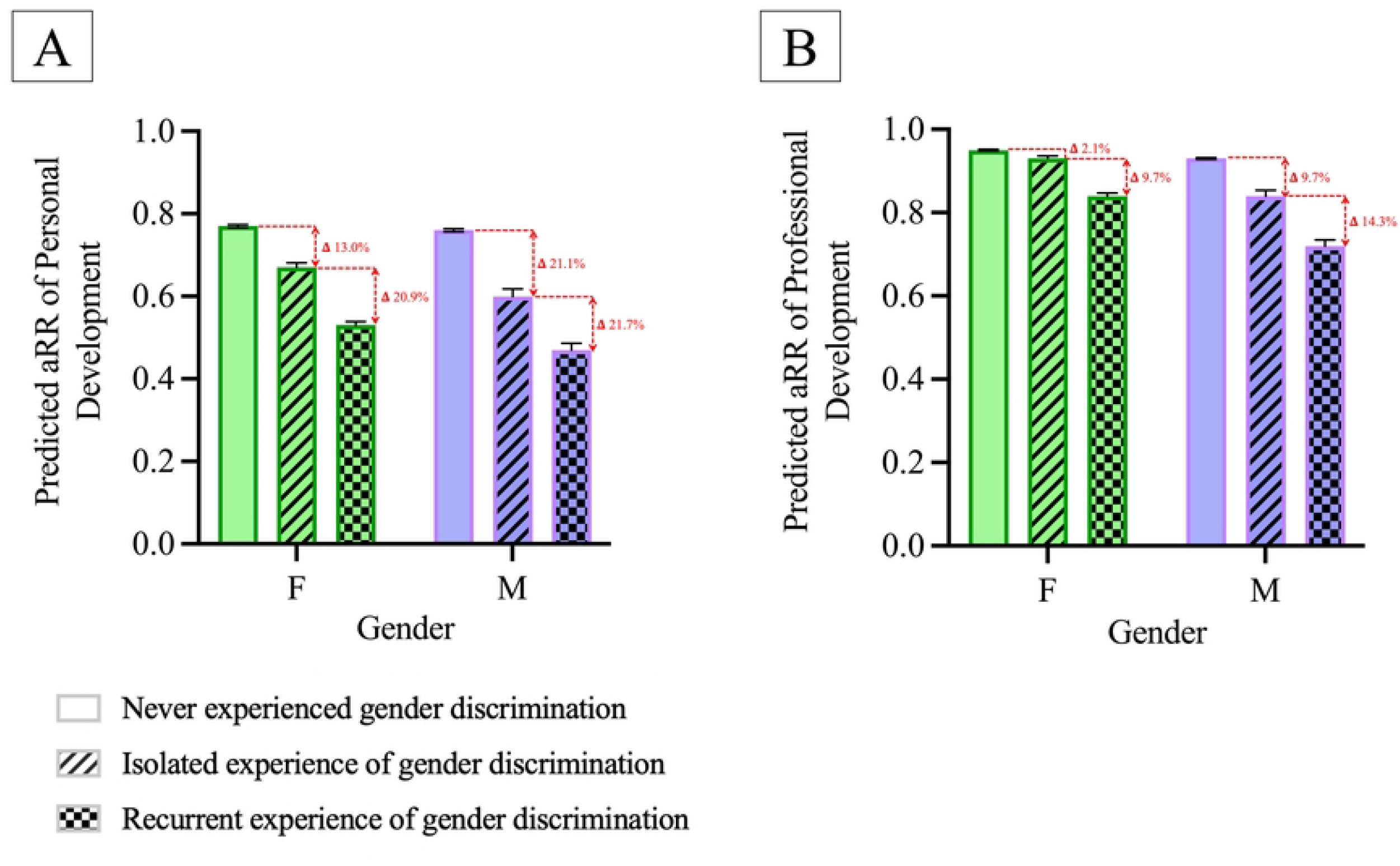
Predicted Personal and Professional Development in Males and Females by Frequency of Gender Discrimination. Panel A illustrates the predicted adjusted relative risk (aRR) of personal development for female (green) and male (purple) medical students based on their experience of gender discrimination (never, isolated, or recurrent). Panel B shows the predicted aRR of professional development for female (green) and male (purple) medical students by their experience of gender discrimination (never, isolated, or recurrent). For both panels, the red arrows and corresponding delta (Δ) values represent the percentage decrease in aRR as the frequency of reported gender discrimination increases from no discrimination to isolated discrimination, and from isolated to recurrent discrimination, for both genders. All predicted relative risks are adjusted for race/ethnicity and childhood household income.

The change in predicted relative risk of the school fostering professional development among students who never experienced gender discrimination compared to students who experienced isolated discrimination is significantly higher in males than females (exp(b)=0.93, 95% CI 0.92-0.94, p<0.001, ***Figure 4B***). The change in predicted relative risk of the school fostering professional development among students who experienced isolated gender discrimination compared to students who experienced recurrent discrimination is significantly higher in males than females (exp(b)=0.95, 95% CI 0.93-0.97, p<0.001).

## Discussion

This study uncovers significant, progressive, and consistent links between experiences of gender discrimination and the lack of personal and professional development fostered by medical schools among their students. Although previous research has highlighted the existence of gender discrimination in undergraduate medical education(5,10,12), this is the first study to investigate its influence on shaping medical student PPIF.

### Greater gender discrimination experienced by female students

As expected, our study indicates that female students report recurrent and isolated experiences of gender discrimination more frequently than their male counterparts. Women often face more adverse experiences in men-dominated fields like medicine, including stereotyping and bias(13,14), and disparaging comments related to their gender, gender discrimination, and sexual harassment.(15) These experiences can be attributed to societal and institutional norms that have historically favored men in medical professions.(14) Critically, over one-third of female medical students in our study experienced gender discrimination and over one-fifth experienced it recurrently. These frequent experiences of discrimination have been attributed to diminished career aspirations among women medical trainees(6), as well as negative impacts on self-esteem(16), self-efficacy(17) and mental health(18), likely impeding their PPIF. Not only do female students face significantly more gender discrimination than males, but this inequity persists into their careers, as evidenced by women faculty who also report higher rates of discrimination compared to their male counterparts.(19) Possibly due to the continued experience of gender discrimination, women faculty experience lower career satisfaction,(20,21) higher burnout rates,(22) less research productivity,(15) lower earnings(15) and slower career advancement,(15)and hold far fewer full professorships (29%) and departmental chair positions (24%).(23) Similarly, among women medical students, gender discrimination may impede PPIF by contributing to burnout(24), and diminishing opportunities for leadership and professional growth.(6) Addressing gender discrimination early in medical education might help mitigate the long-term impacts on female students.

### Gender disparities in medical student development

The AAMC identifies personal and professional development as key competencies in medical education.(1) Our study indicates that both women and men are equally likely to attest that their schools fostered their professional development, possibly reflecting an appropriately increasing focus on gender inclusivity in medical education in recent times.(25) Women reporting similar rates of professional development despite experiencing higher rates discrimination perhaps suggests that women may employ resilience and coping strategies that buffer against the adverse effects of gender discrimination on professional aspirations.(26) Our study also suggests that female medical students perceive a lack of support for their personal development from their medical schools compared to their male counterparts. This discrepancy may stem from underlying systemic biases and gender-specific challenges that women face in medical education.(12) Studies have shown that the gender discrimination and microaggressions that female students often encounter in the learning environment can hinder their sense of belonging and support within the academic environment.(5,12)

### Influence of gender discrimination on personal and professional development among female and male students

With progressively greater levels of gender discrimination, all students report lower PPIF, in our study. Gender discrimination in educational environments has been shown to negatively impact educational outcomes for both women and men(27), contributing to decreased academic performance(28), and biased evaluations.(27) Interestingly, our study observes that gender discrimination is associated with a greater negative influence on PPIF in men compared to women. This finding aligns with other research reporting a greater influence of gender discrimination on career decisions among men than women, despite fewer men experiencing gender discrimination, suggesting that men may weigh experiences of discrimination more heavily in their career decisions.(6) This may be because men experience gender discrimination as a more significant violation of social norms, which could lead to greater psychological impact and disruption in their development process.(29) This is consistent with the concept of stereotype threat, where being in the minority or facing unexpected social discrimination can erode self-identity and performance.(30) Men may have fewer coping mechanisms for managing gender discrimination than women, who, due to more frequent exposure, may be more resilient, potentially exacerbating gender discrimination’s negative effects on the PPIF of men. It is also possible that the gender discrimination experienced by men is qualitatively different, perhaps due to fewer support avenues available for men in medicine to discuss these experiences compared to women.

### Gender discrimination’s greater influence on personal than professional development

Among both women and men, gender discrimination appears to impact personal development more severely than professional development. This difference may be due to the explicit focus of medical education on developing clinical competence and professional behaviors, as delineated by the Liaison Committee on Medical Education (LCME). Notably, the LCME accreditation guidelines do not explicitly address personal development. Including this in their standards could serve as another intervention to enhance holistic PPIF among students facing gender discrimination. Given the influence of gender discrimination on multiple factors influencing personal development, including feelings of isolation, professional self-confidence and satisfaction, self-esteem, collegiality,(31) and specialty choice and residency program selection(6), medical schools should consider specific efforts to foster student personal development among students experiencing gender discrimination.

## Limitations

Our binary categorization of participants as either women or men excludes non-binary and other gender identities, potentially overlooking their unique experiences of gender discrimination and its impact on educational outcomes. Similarly, the absence of sexual orientation data limits our ability to explore how intersecting identities, such as sexual minority status, may influence personal and professional development. In addition, high non-response rates for sensitive variables, such as childhood household income and experience of gender discrimination, required us to rely on imputation. While multiple imputation slightly shifted the gender distribution from a female to a male majority, the consistency of results between the complete case and imputed analyses supports the robustness of the findings. Furthermore, the GQ captures only a subset of gender discrimination experiences; however, those included are significant and can have serious psychological and professional impacts. While the GQ measures used as proxies for personal and professional development assess institutional success in fostering student growth—perhaps appropriately attributing responsibility to the school—they may not fully capture the extent of development, or lack thereof, experienced by students. We recognize that the multi-domain and dynamic nature of identity development during medical training likely extends beyond the development fostered by the institution to also include individual factors, though institutional efforts remain crucial.(2,4)

## Implications

Our findings highlight a persisting need for systemic reforms in medical education to address gender discrimination that is experienced by both female and male students, and to further investigate how gender discrimination may influence educational outcomes among gender-diverse students. Addressing gender discrimination in the learning environment, as well as racial discrimination, as our previous work suggests, may serve to promote holistic development for all students.(32) We propose considering the integration of PPIF as a critical equity metric and formally including PPIF parity for students of all racial, ethnic, and gender backgrounds in the LCME accreditation standards. Educators, institutions, and organizations like the LCME should consider implementing initiatives to combat gender bias, benefiting not only students who are women but also enhancing the development of students who are men, who experience more significant declines in PPIF with increased gender discrimination. Fostering a sense of belonging in medicine by addressing unconscious biases and transforming academic medicine environments to recognize, hear, and value the contributions of diverse learners can positively influence PPIF in medical school.(33,34) Validated tools that can systematically measure and help achieve a climate of equity and inclusion within the medical school learning environment, such as the PRODIGIE tool developed by our team, may provide medical schools with actionable insights to foster equity and inclusion,(35) ultimately improving educational outcomes, including PPIF, for all students.

## Conclusion

Our study provides empirical evidence highlighting the influence of gender discrimination on the effectiveness of medical schools in nurturing students’ PPIF. Female students experience gender discrimination more frequently than their male counterparts. Women were also less likely to report that their medical school fostered their personal development than men. Increasing frequency of gender discrimination correlates with a decreased likelihood, for both women and men, that their school supported their PPIF. However, men experienced sharper declines in PPIF rates with increasing frequency of gender discrimination compared to women. Overall, these findings highlight a potential influence of gender discrimination on development within educational settings, calling for targeted interventions to address gender discrimination in medical school and foster an environment conducive to equitable development for all students.

## Data Availability

The data underlying the results presented in the study are available on request from the Association of American Medical Colleges (AAMC).

## Acknowledgements

This work is supported by the National Institute of Health: NIGMS grant T32GM136651 (MN), NIAID grant F30AI157227 (MN), NIGMS grant R35GM153263 (DB). The funder had no role in the design and conduct of the study; collection, management, analysis, and interpretation of the data; preparation, review, or approval of the manuscript; and decision to submit the manuscript for publication. This material is based upon data provided by the Association of American Medical Colleges (“AAMC”). The views expressed herein are those of the authors and do not necessarily reflect the position or policy of the AAMC.

## Conflict of Interest Statement

Alexis Webber, Mytien Nguyen, Hyacinth RC Mason, Mayur M. Desai, and Shruthi Venkataraman report no conflicts of interest. Dowin Boatright received NIH grant (R35GM153263) support. Alexandra Hajduk received NIH grants (R01 GM146147, R01 MD018928, R01 HL160822) and Burroughs-Welcome Fund travel support. Mytien received NIGMS (T32GM136651) and NIAID (F30AI157227) grant support. Sarwat Chaudhry received support from NIH/DHHS (1R01MD018928-01A1) and NIH/NIGMS (GM146147). Tonya L. Fancher received grants and consulting fees from the American Medical Association, honoraria from Montefiore School of Medicine, Yale School of Medicine, University of Minnesota School of Medicine, travel support from ACGME, and holds a leadership role with HRSA.

---

**Table 1:** Participant Demographic Characteristics.

| Characteristic | Original Dataset<br>No. (%)<sup>1</sup><br>N = 21 728 | Imputed Dataset<br>No. (%)<br>N = 37 610 |
| --- | --- | --- |
| Gender |  |  |
| Female | 11 035 (50.8) | 18 200 (48.4) |
| Race/Ethnicity |  |  |
| African American or Black | 1 277 (5.9) | 2 460 (6.5) |
| Asian | 4 320 (19.9) | 7 810 (20.8) |
| Hispanic | 1 458 (6.7) | 2 438 (6.5) |
| multiracial | 1 349 (6.2) | 2 410 (6.4) |
| other<sup>2</sup> | 576 (2.6) | 1 137 (3.0) |
| white | 12 748 (58.7) | 21 355 (56.8) |
| Yearly Childhood Household Income |  |  |
| <\$50,000 | 4 229 (19.4) | 7 983 (21.2) |
| \$50,000 to <\$75,000 | 3 623 (16.7) | 6 321 (16.8) |
| \$75,000 to <\$125,000 | 6 648 (30.6) | 11 184 (29.7) |
| \$125,000 to <\$200,000 | 3 621 (16.7) | 6 075 (16.2) |
| ≥ \$200,000 | 3 607 (16.6) | 6 047 (16.1) |
| Experience of Gender Discrimination |  |  |
| Never | 17 068 (78.5) | 29 622 (78.8) |
| Isolated | 1 824 (8.4) | 3 129 (8.3) |
| Recurrent | 2 836 (13.1) | 4 859 (12.9) |
| Personal Development |  |  |
| Yes | 15 920 (73.3) | 27 195 (72.3) |
| Professional Development |  |  |
| Yes | 20 074 (92.4) | 34 488 (91.7) |

<sup>1</sup>Only data for complete cases included in table to facilitate comparisons with imputed dataset. Missing data for demographic variables from the original sample size of 37,610 as follows: race/ethnicity (N=2,429), yearly childhood household income (N=8,436), experience of gender discrimination (N=7,917), personal development (N=7750), professional development (N=7459).
<sup>2</sup>American Indian/Alaska Native and Hawaiian Native/Pacific Islander (HNPI) students were included as part of the 'other' group, due to their small sample size.

## Notes

### Competing Interest Statement

The authors have declared no competing interest.

### Funding Statement

Yes

### Author Declarations

Using fully anonymized student-level data from the AAMC data warehouse, we conducted a retrospective cross-sectional study of 40,971 Doctor of Medicine (MD) matriculants from the 2014-2015 and 2015-2016 academic years who graduated between 2016 and 2020 and took the American Medical College Application Service (AMCAS) survey and Graduation Questionnaire (GQ). Students provided explicit electronic consent on AMCAS by actively selecting an option to allow the use of their data for research purposes. Participation in the GQ was voluntary, and students were informed that their responses could be used for research purposes, providing implied informed consent by choosing to participate. The Yale School of Medicine institutional review board waived the need for additional ethics review and informed consent, as all data were fully deidentified by the AAMC prior to transfer and researcher access. Data were accessed for research purposes between February 1, 2024, and June 3, 2024.

